# The genomic and evolutionary landscapes of anaplastic thyroid carcinoma

**DOI:** 10.1101/2023.04.10.23288365

**Authors:** Peter Y.F. Zeng, Stephenie D. Prokopec, Stephen Y. Lai, Nicole Pinto, Michelle A. Chan-Seng-Yue, Roderick Clifton-Bligh, Michelle D. Williams, Christopher J. Howlett, Paul Plantinga, Matthew Cecchini, Alfred K. Lam, Iram Siddiqui, Jianxin Wang, Ren X. Sun, John D. Watson, Reju Korah, Tobias Carling, Nishant Agrawal, Nicole Cipriani, Douglas Ball, Barry Nelkin, Lisa M. Rooper, Justin A. Bishop, Cathie Garnis, Ken Berean, Norman G. Nicolson, Paul Weinberger, Ying C. Henderson, Christopher M. Lalansingh, Mao Tian, Takafumi N. Yamaguchi, Julie Livingstone, Adriana Salcedo, Krupal Patel, Frederick Vizeacoumar, Alessandro Datti, Liu Xi, Yuri E. Nikiforov, Robert Smallridge, John A. Copland, Laura A. Marlow, Martin D. Hyrcza, Leigh Delbridge, Stan Sidhu, Mark Sywak, Bruce Robinson, Kevin Fung, Farhad Ghasemi, Keith Kwan, S. Danielle MacNeil, Adrian Mendez, David A. Palma, Mohammed I. Khan, Mushfiq Shaikh, Kara M. Ruicci, Bret Wehrli, Eric Winquist, John Yoo, Joe S. Mymryk, James W. Rocco, David Wheeler, Steve Scherer, Thomas J. Giordano, John W. Barrett, William C. Faquin, Anthony J. Gill, Gary Clayman, Paul C. Boutros, Anthony C. Nichols

## Abstract

Anaplastic thyroid carcinoma is arguably the most lethal human malignancy. It often co-occurs with differentiated thyroid cancers, yet the molecular origins of its aggressivity are unknown. We sequenced tumor DNA from 329 regions of thyroid cancer, including 213 from patients with primary anaplastic thyroid carcinomas and multi-region whole-genome sequencing. Anaplastic thyroid carcinomas have a higher burden of mutations than other thyroid cancers, with distinct mutational signatures and molecular subtypes. Specific cancer driver genes are mutated in anaplastic and differentiated thyroid carcinomas, even those arising in a single patient. We unambiguously demonstrate that anaplastic thyroid carcinomas share a genomic origin with co-occurring differentiated carcinomas, and emerge from a common malignant field through acquisition of characteristic clonal driver mutations.

## Statement of Significance

Anaplastic Thyroid Cancer is the single most lethal human cancer. Surprisingly, it often evolves alongside a highly non-lethal form of differentiated thyroid cancer. demonstrate how these two diseases evolve from a common ancestor, leading to differential evolutionary trajectories and therapeutic targets.

## Introduction

Human malignancies range widely in their lethality. Some tumor types, like differentiated thyroid carcinoma (DTC) and prostatic adenocarcinomas, are typically indolent with long life histories and managed with minimally-invasive therapies or even surveillance protocols^1,2^. Others, like pancreatic adenocarcinomas, progress rapidly after initial diagnosis and are refractory to most therapeutic interventions^3^. This variability between tumor types is mirrored within them as well, leading to the development of prognostic tests that predict tumor aggressivity from transcriptomic or proteomic features^4–6^.

At one extreme of this spectrum of tumor lethality lies anaplastic thyroid carcinoma (ATC). ATC is arguably the most lethal human tumor type, with a median survival of ∼12 weeks: some patients succumb to their disease within days of diagnosis^7,8^. ATC usually presents dramatically, with rapid onset of airway and oesophageal obstruction due to explosively growing neck masses^7,8^. Primary ATCs are frequently surgically inoperable due to encasement of nerves, blood vessels and the airway. Radio-and chemo-resistance are common, and distant metastases are near-ubiquitous^7,9^. ATC is a major clinical dilemma and a model for understanding lethal, treatment-resistant cancer.

The aggressiveness of ATC is particularly intriguing because of its life history: ∼80% of ATCs occur in the context of a prior history of thyroid cancer or with a distinct co-occuring region of DTC (co-DTC), most frequently papillary thyroid cancer (PTC) ^10–14^. Thus intriguingly, the most lethal human malignancy frequently arises in the context of one of the most indolent. ATC presents with undifferentiated pathological features, suggesting that at least a subset may evolve *via* dedifferentiation of DTCs: either well-or poorly-differentiated. There has been little molecular confirmation of progression from a well to poorly differentiated tumor, nor testing of the mutational field effect surrounding lethal ATC. Several studies have sequenced small numbers of ATCs with limited genome coverage, highlighting frequent *TP53* mutations and a subset of tumors with multiple oncogene driver mutations^15–23^. However, the genomic landscape of ATC remains largely unknown, and the molecular characteristics of their evolutionary relationship to DTC remain elusive.

To fill these gaps in our understanding of ATC pathobiology, we established a 15-site consortium called the Global Anaplastic Thyroid Cancer Initiative (GATCI). We report the mutational landscape of 329 thyroid carcinomas, including 179 primary ATCs and 34 co-occurring regions of DTC within ATCs. In order to identify genomic alterations enriched in ATC or its co-DTC region, we sequenced 115 papillary thyroid regions without co-occuring ATC from 112 patients. These data reveal distinct genomic subtypes of ATC, with elevated mutational density and signatures. ATCs harbour a distinct set of driver mutations from DTCs, including multiple recurrent tumor suppressor hits. ATC arising in the context of DTCs share a common evolutionary origin, but acquire additional specific driver mutations, some of which are recurrently altered only in anaplastic carcinoma components.

## Results

### Cohort summary

We collected 329 thyroid cancer regions from 291 patients, including 179 regions of primary ATC and 115 regions of PTC. A detailed breakdown of samples is available in **Supplementary Figure 1** and **Supplementary Table 1a-c**. These tumor samples (and in most cases, a blood reference sample) were characterized for copy number aberrations (CNAs) by SNP microarrays. Single nucleotide variants (SNVs) were identified by exome (WXS) or whole-genome sequencing (WGS), along with RNA-sequencing (RNA-seq) when open biopsies provided sufficient material. Deep targeted sequencing was used for validation, and nested PCR to detect TERT promoter mutations (**Supplementary Table 1**). ATCs were sequenced to a depth of 123 ± 14x and reference samples to 116 ± 13x (median ± standard-deviation, SD) by WXS and 33 ± 4x or 34 ± 4x, respectively by WGS. Reads were aligned and somatic variant detection performed using a validated pipeline^6,24^. Recognizing that ATCs can have low tumor cellularity, we validated 1,140 candidate mutations in 54 ATCs by deep targeted-sequencing on an orthogonal platform sequencing platform to with median F1 of 0.914 (**Supplementary Figure 2**). In contrast to the strong female preponderance of PTC in the Cancer Genome Atlas (TCGA) (F:M 2.93:1^25^), our ATC cohort was sex-balanced (F:M 1.11:1), consistent with previous reports^17^. Median age at diagnosis was ∼70 years for ATCs and ∼50 years for PTC within our cohort. Most ATC patients presented with locally advanced disease: 87% with T4b disease and 70% with nodal involvement. Distant metastases were common (43%), and median survival was 126 days from diagnosis. PTCs within the GATCI project were equally distributed by tumor extent (T1-4), and all tumors were surgically resected, with median follow-up of 10.9 years. Tumors were subjected to consensus pathology review and manually macro-dissected to maximize cellularity (**Supplementary Table 1**).

### ATCs are moderate mutation burden CNA-type cancers

While differentiated thyroid cancers (DTCs) have fewer somatic mutations of all types than almost any other cancer type^25^, ATCs have been suggested to be highly mutated, particularly by CNAs^17,26^.

In this large genome-wide ATC cohort we identified 3.8 ± 1.2 SNVs/Mbp of DNA sequenced (**Figure 1A (top) and 1C**). These were accompanied by 120 ± 44 CNAs (mean ± 99% CI; median = 83.5; **Figure 1B,D**). The average CNA was 6.21 ± 1.33 Mbp (mean ± 99% CI; median = 4.95 Mbp). Relative to 32 tumor types^27^, ATCs show both more CNAs (**Figure 1D**) and more SNVs/Mbp (**Figure 1C**) than PTCs, but fewer than most other adult cancer types. Despite their clinical aggressiveness, they are not amongst the most highly mutated tumor types, nor did they show atypically high inter-tumor variability relative to other cancer types.

**Figure 1:**
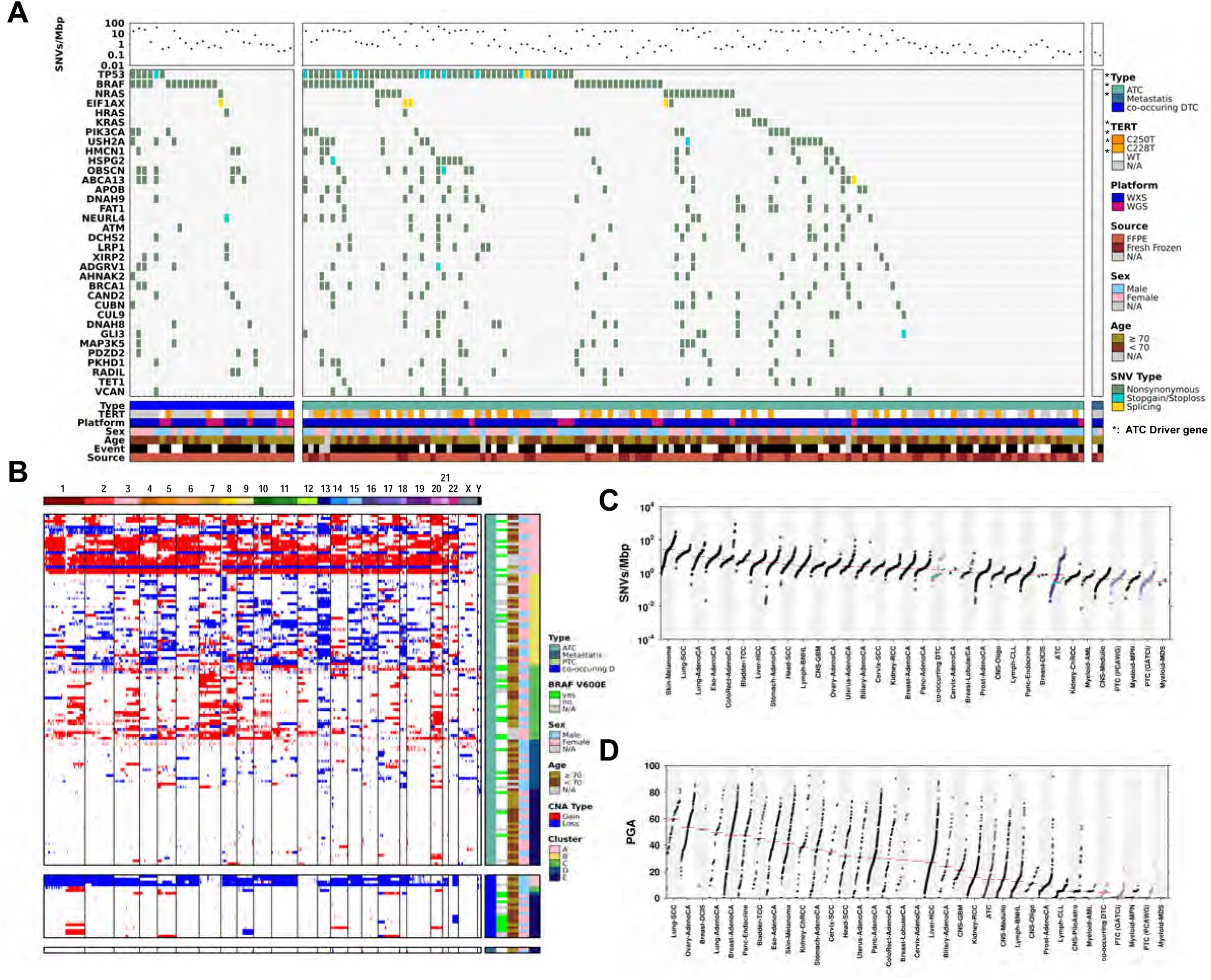
Somatic Mutational Landscape of Anaplastic Thyroid Cancer. **A**) Anaplastic thyroid cancers (ATCs) vary significantly in in mutation density (single nucleotide variants [SNVs] per million bp [Mbp] of DNA covered). **B)** Copy number aberrations (CNAs) across ATC; consensus clustering was used to identify the optimal method and designation for sample groupings. Each row represents the CNA profile for a single undifferentiated tumor along the genome (chromosome 1 on the left to chromosome Y on the right), ordered by percent genome altered (PGA) within each subtype. Metrics of mutation density for ATC, co-occurring DTC and pappilary thyroid cancer (PTC) were compared with 32 additional tumor types available in the PCAWG (Pan-Cancer Analysis of Whole Genomes) dataset: **C)** SNVs/Mbp and **D)** total PGA. Light purple indicates the PCAWG thyroid carcinoma cohort (primarily PTC) and similar GATCI PTC cohort; co-occurring DTC samples are shown in medium purple while ATC are in dark purple; for SNVs/Mbp, x’s indicate samples without a matched normal – these typically have higher than average rates. PGA for the GATCI PTC samples is similar to that of the PCAWG thyroid carcinoma cohort. Blue points for SNVs/Mbp indicate results from WGS cohort. ATC shows higher PGA than either co-occurring DTC or PTC, and a lower rate of point mutations than co-occurring DTC.

Seven driver genes were affected by recurrent SNVs in ATC *via* SeqSig driver gene analysis^6^ (false discovery rate [FDR] < 0.01), including *TP53*, *BRAF*, *NRAS* and *PIK3CA* (**Figure 1A**, **Supplementary Figure 3A**). Non-functional or partially functional^28^ non-synonymous SNVs in *TP53* were associated with elevated mRNA abundance (Wilcoxon rank sum test p = 0.0067, **Supplementary Figure 3B**, **Supplementary Table 2**). Another of these drivers, *HMCN1*, was expressed only in ATC but not in normal thyroid tissue (transcript per million [TPM] > 1 in more than 20% of profiled normal tissues). Several mutations previously described in ATC were detected at low frequency – *EIF1AX* mutations were detected in five tumors, including three with the p.A113X splice-site mutation reported to be exclusive to thyroid cancers^17^ and two missense mutations (G9R). These tended to co-occur with *RAS* mutations, consistent with previous reports (**Supplementary Figure 4A**)^17^. *BRAF*^V600E^ and *RAS* mutations were mutually exclusive, while *BRAF*^V600E^ and *PIK3CA* mutations co-occurred^15,17^ (hypergeometric test p=0.0052; **Supplementary Figure 4A**). Promoter mutations upstream of *TERT* were correlated with *BRAF* mutations (χ^2^ p = 0.008).

We assessed germline variants in known cancer pre-disposition genes^29^. Ten genes with heterozygous germline SNVs in at least 2 patients (2.4% of ATCs), with a maximum recurrence of four patients (5%) for *RECQL4* (**Supplementary Table 3**). This included multiple DNA damage repair genes, for example *BRCA2* and *FANCF*. Given the lack of strong recurrence however, much larger cohorts will be required to understand the germline correlates, if any, of ATC. We therefore focused on somatic mutations.

Individual ATCs varied significantly in their underlying mutational processes, with activation COSMIC signatures 1, 2, 5, 6, 13 and a rare novel signature (**Figure 2A**). The latter was characterized by G[T>G]G mutations and was strongly detected in only three tumors (**Figure 2B**). It may embody unknown sequencing artefacts or an as-yet-undescribed mutational process. Other types of thyroid cancers have been reported to harbour activation of COSMIC signatures 1, 2, 5 and 13^30,31^, and ATC mirrors these. COSMIC 5 describes a diverse range of point mutations consisting of low frequency pyrimidine transition mutations, and COSMIC signature 13 is attributed to AID/APOBEC activity^30^, but surprisingly none of these signatures were associated with sex, age, CNA subtype, *TERT* promoter status or overall survival (ANOVA, FDR > 0.1). Thus ATC harbours more somatic SNVs than other thyroid cancers, but fewer than other cancer types, with no widely recurrent driver mutations or mutational processes.

**Figure 2:**
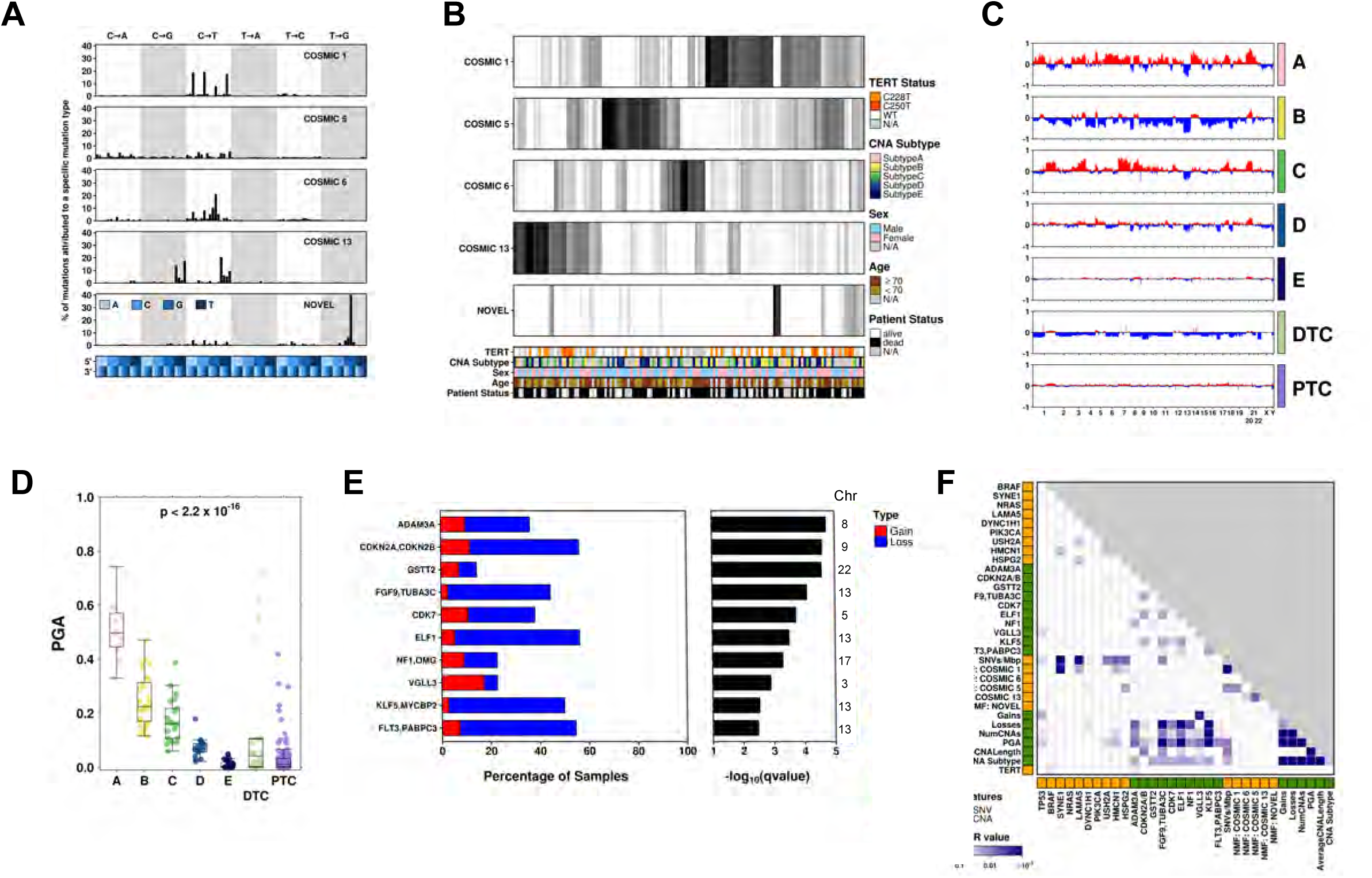
Genomic Features of ATC and Their Associations with Clinical Features. **A**) NMF identified five trinucleotide signatures within ATC (n = 132 tumors), four of which matched known COSMIC signatures and one novel signature. Within each signature, the percent of mutations within the cohort presenting each base change (broken down by trinucleotide context); **B)** for each patient, the proportion of SNVs that contribute to each signature. There are no associations between these signatures and covariates shown (Spearman’s correlation p-value < 0.1). **C)** Average CNA profiles and **D)** distribution of PGA for samples from each ATC subtype, co-occurring DTC and PTC, showing a significant difference between groups (one-way ANOVA, p < 0.01). **E)** GISTIC was used to identify recurrent CNAs within ATC. **F)** Pearson’s χ2, Spearman’s correlation, Wilcox or Kruskal-Wallis tests were used to assess overlap across genomic features in ATC (recurrently altered genes by SNV or CNAs, SNVs/Mbp, PGA, trinucleotide signatures, CNA metrics); shading indicates FDR-adjusted p-value.

By contrast, ATC harboured a background of multiple recurrent chromosome-scale events, including loss of chromosome 13 (39% of tumors), gain of chromosome 7 (26% of tumors) and gain of chromosome 20 (35% of tumors). Consensus clustering revealed that these large-scale changes reflect five distinct CNA subtypes, denoted A-E (**Figure 1B, 2C**). Each subtype is defined by characteristic genomic abnormalities, such as wide-scale amplifications with occasional arm-level deletions for Subtype A (11% of ATCs) and large-scale deletions with cases of uniparental polysomy, similar to Hürthle cell thyroid cancers (HTC)^32,33^, for Subtype B (29%). Subtype C (20%) is dominated by gains of chromosomes 7 and 20 (similar to HTC^32,33^) and 1q amplifications (similar to PTC^25^), while Subtype D (15.5%) demonstrates many focal regions of CNA. Subtype E (24.5%) shows a quiet copy number profile, analogous to similar profiles in breast, prostate and head and neck cancers^34–36^. These subtypes were associated with overall genomic instability (**Figure 1D**), age, and TP53 mutation status (**Supplementary Table 4**), but were independent of tumor cellularity and ploidy (**Supplementary Figure 4B–H**). These broad changes were accompanied by many highly-recurrent focal driver CNAs (**Figure 2E**, q < 0.01). Loss of *CDKN2A* was wide-spread (42% of ATCs)^17,37^, as was loss of *BRCA2* (33.6% of ATCs). Other prominent drivers were loss of a 62 kbp region on 22q containing *GSTT2* and *GSTTP1*, loss of regions on chromosome 13 involving *FGF9*, *MYCBP2* and *FLT3* and a 429 kbp gain on chromosome 3p containing *VGLL3*. Many of these recurrent CNAs were associated with mRNA changes (**Supplementary Table 2**, **Supplementary Figure 4I–K**).

We next sought to identify fusion genes using RNA-sequencing data, reasoning that these are common in other thyroid cancers and often attractive therapeutic targets. We identified 84 fusion genes in primary ATC using fusioncatcher^38^, covering 144 total partner genes, and none previously reported in COSMIC (**Supplementary Table 5A**). The typical ATC harboured a median of 3 distinct fusion genes, with 25% of samples containing 6 or more fusions and two harbouring a maximum of 11 fusions. Only five fusion genes occurred in two or more patients, including *LINC01133*:*SAMHD1*, *MDM4:TRA* and *KDSR*:*ANAPC7*. Fusions involving *FN1* and *COL1A1* were associated with higher than average abundance of these genes. For *FN1*, this presented as 4.3x higher TPM in tumors with an *FN1* fusion (Wilcoxon p = 0.007) and for *COL1A1*, fusions corresponded to 12x higher TPM (one sample; **Supplementary Table 5B**). Increased abundance of both genes has previously been associated with tumor cell migration and invasion in multiple tumor types^39,40^.

To identify co-occuring mutation processes underlying ATC pathogenesis, we analyzed inter-associations of mutation-density, mutation-signature and driver-gene features (**Figure 2F**). Multiple associations were detected, including a strong correlation between SNV mutation density (SNVs/Mbp sequenced) and COSMIC 1 signature, *SYNE1* and *LAMA5* mutations (FDR < 10^-3^). Trinucleotide signatures were not correlated to CNA subtypes (p > 0.05; one-way ANOVA; **Supplementary Figure 4L**), suggesting the processes driving CNA diversification is independent from those driving SNV patterns.

### Clinico-Genomics of ATC

To assess the clinical relevance of these somatic mutational features, we first assessed the impact of clinical factors on overall survival (**Supplementary Table 1**). Treatment with radiotherapy (FDR = 1.7 x 10^-5^) or surgery (FDR = 0.0089) were positively associated with patient outcome, as previously described^41^. Nodal involvement (FDR = 0.009), distal metastasis (FDR = 0.025), leukocytosis (FDR = 0.056) and patient age (FDR = 0.08) were all associated with reduced survival. The use of surgery and patient age strongly stratified ATC survival outcomes (**Supplementary Figure 3A**).

We then considered the association of individual driver features with patient outcome, including patient age and treatment as covariates in time-to-event analyses. No genes with point mutations in ≥5% of ATCs were associated with overall survival (FDR > 0.1, **Supplementary Table 6**). Although CNA subtypes A-E were not associated with overall survival (p = 0.6, **Supplementary Figure 3B**), tumors with fewer CNAs more often presented with distant metastases (Wilcoxon rank sum test: FDR = 0.034; median percent genome altered [PGA] for patients with and without distant metastases = 8.6% and 11.6% respectively). PGA was not associated with overall survival (median dichotomized; HR = 0.93 [95%CI, 0.59-1.47], p = 0.77, adjusted for patient age and surgery), unlike in several other tumor types^35,42,43^. *BRCA2* deletion was surprisingly associated with better survival (HR = 0.48 [95%CI, 0.29-0.80], p = 0.005; **Supplementary Figure 3D**), but no other recurrent CNA was (**Supplementary Table 7**).

By contrast, CNA subtypes showed distinctive clinical hallmarks: subtype A was enriched for older patients with better overall survival, subtype B was depleted for patients with nodal involvement as well as moderate enrichment for patient death, subtype C was enriched for younger patients, subtype D was enriched for male patients with metastatic disease and subtype E showed no associations with clinical variables (**Supplementary Figure 3E**). Despite limited statistical power, patients with a lower mutation rate (>10 SNVs/Mbp) had significantly better survival (HR = 0.51, [95%CI, 0.33-0.77], p = 0.002; **Supplementary Figure 3F**). Thus the mutational features and subtypes of ATC are associated with divergent clinical presentation and outcome.

### ATC and DTC evolve in parallel from a mutagenic field

It has been suggested that a subset of ATCs evolve from pre-existing DTCs^11-13,23,44,45^. Given the strong association of ATC CNAs with clinical and molecular phenotypes, we first focused on copy number changes. We compared SNV and CNA profiles between PTCs, co-DTCs and ATCs using both newly-profiled PTC samples from this study and the TCGA dataset^25^. The two recurrent arm-level events of PTC were 1q amplification and 22q deletion^25^, both of which occurred at similar frequencies in both ATCs (**Figure 1B, 1H**) and the co-occurring DTC components of ATCs (**Figure 1B, 3C**). All recurrent focal CNAs in PTC were also detected in ATC, but several CNA drivers were significantly more frequent in ATC (**Figure 3B**). Loss of *CDKN2A* was recurrent in both co-occurring DTCs (**Figure 3C**) and ATC, but was rare in PTCs (∼5%). *BRCA2* was also frequently deleted in ATCs (33.6%), uncommon in DTCs (13.6%) and rare in PTCs (4.5%), as was *RB1* which is on the same chromosome arm. Several other regions showed higher rates of copy number change in ATCs and co-occurring DTCs, including a broad region on chromosome 20q harboring 328 genes was preferentially amplified in ATC relative to both cohorts of PTCs (proportion-test, FDR < 0.01; **Supplementary Table 8**). This region harbors several cancer driver genes, for example *ARFGEF2*, *CHD6* and *GNAS*, all ubiquitously expressed in thyroid tissue^46^.

**Figure 3:**
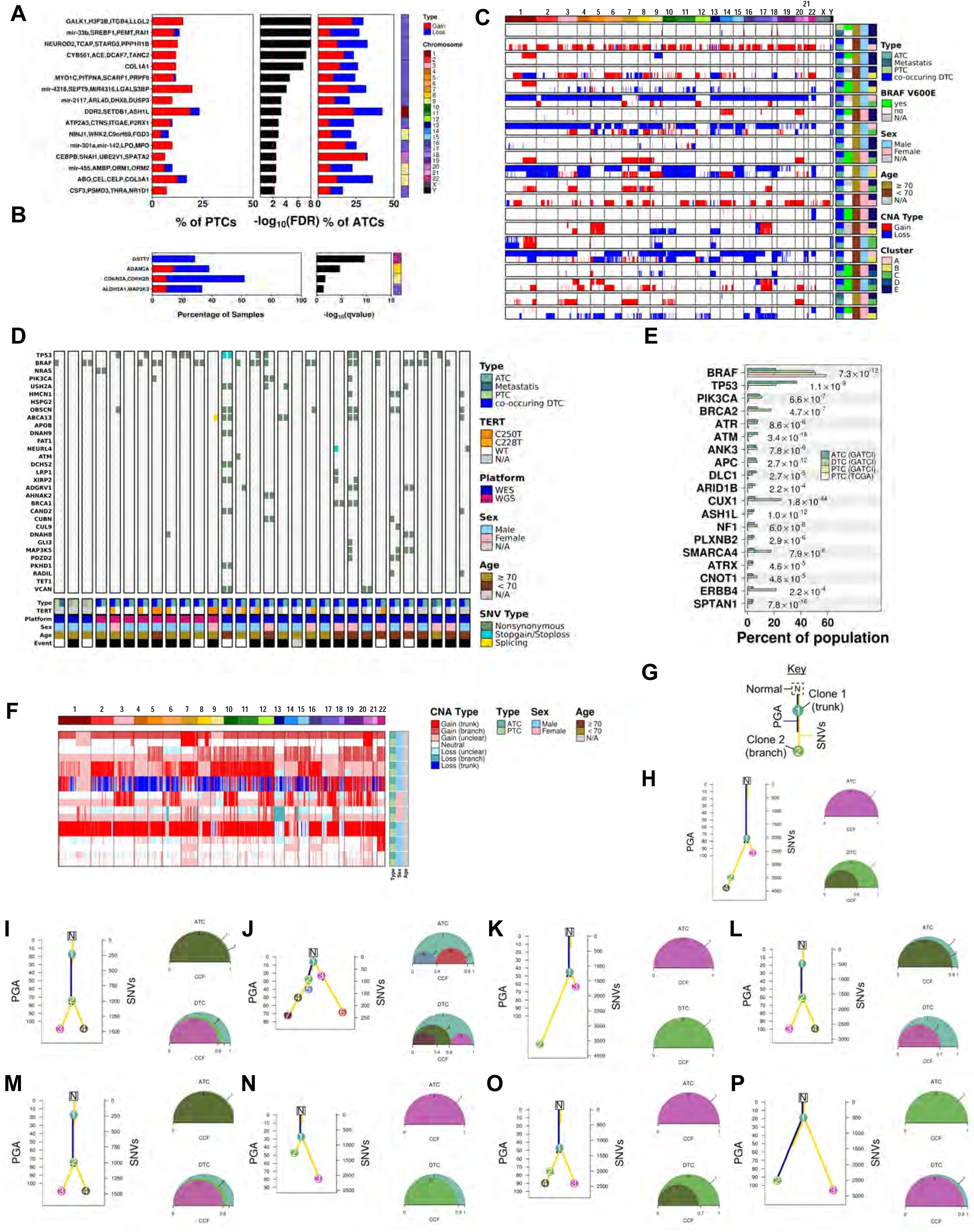
ATC and co-occuring DTC Share Clonal Origins, But With Distinct Driver Events. **A**) GISTIC was used to identify recurrent CNAs within co-occurring DTC. Copy number losses of *CDKN2A* were more frequent in ATC than co-occurring well/poorly differentiated samples (DTC). **B)** GISTIC was used to identify recurrent CNAs within co-occurring DTC. Copy number losses of CDKN2A were more frequent in ATC than co-occurring well/poorly differentiated samples (DTC). **C)** Twenty-one patients had CNA profiles generated through copy-number arrays for multiple tumor regions. Each plot shows the genomic CNA profiles for a single patient, with different tumor regions as rows. Covariates on the right indicate ATC subtype (where available), tumor type (ATC, co-occurring differentiated thyroid cancer [DTC] or metastasis), patient sex, patient age at diagnosis and *BRAF* V600E status. **D)** 30 patients had WXS or WGS in multiple regions. Each subplot show the mutational profile of the most frequently mutated genes in the paired samples. **E)** Gene-wise mutation frequencies were contrasted between datasets (GATCI-ATC, GATCI-DTC, GATCI-PTC, and TCGA-PTC) to identify candidate drivers of tumor progression. Genes with a statistically significant difference (proportion-test) between ATC and TCGA-PTC were selected and further filtered to show only known driver genes. False discovery rate (FDR)-adjusted p-values from proportion-tests across the three tumor types (PTC (combined), co-occurring DTC, ATC) are shown. Results for pairwise comparisons are available in **Supplementary Table 3**. **F)** Landscape of Copy number aberrations (CNAs) determined by whole genome sequencing (WGS) for patients with both ATC and paired co-occurring DTC, along with matched normal tissue; CNAs are colour-coded as to their origin (trunk, branch or unclear). **G)** Key for interpreting plots **H)**-**P)**. Left panel: changes in percent genome altered (PGA) are shown by blue lines and corresponds to the left axis while accumulation of SNVs is shown by gold lines and corresponds to the right axis. Right panel: representation of the total cancer cell fraction (CCF) for each distinct subclone for either the ATC (top) or co-occuring DTC (bottom) component. **H-P)** Subclonal reconstruction of the nine patients profiled using WGS, Interestingly, ATCWGS-33 **J)** showed outgrowth of two distinct co-mixed lineages from a mutagenic field.

We next compared the somatic CNA (**Figure 3C**) and SNV profiles (**Figure 3D**) of DTCs co-occurring with ATCs. DTCs co-occurring with ATC showed a larger fraction of their genome affected by CNAs than isolated DTCs (**Figure 3C**). Similarly, Co-occurring DTCs harbored more SNVs than did isolated PTCs or other thyroid cancers arising in individuals without an ATC diagnosis: their overall mutation density was statistically indistinguishable from ATC (4.4 ± 2.8 SNVs/Mbp). Similarly, To explore somatic driver SNVs, we merged the current study with six previous sequencing studies of ATC, PTC and HTC^17,21,23,25,32,33^ (**Supplementary Table 9**). *BRAF* was the only gene more common in differentiated thyroid cancers: it was mutated in 50.9% of PTC, 50.0% of co-occurring DTC and 21.3% of ATCs (**Table 1**, **Figure 3E**). Indeed in matched co-occurring cases, half of all *BRAF*^V600E^ variants were detected solely in the DTC component and not in the ATC component. By contrast multiple other drivers were preferentially mutated in ATCs and co-occurring DTC, including *ATM*, *ATR*, *BRCA2*, *PIK3CA* and *TP53* (**Figure 3E**). As an example, TP53 mutation frequency varied from 0.9% of PTC to 21.4% of co-occurring DTC and 36.8% of ATC (**Supplementary Table 9**). We further compared variant allele frequencies (VAFs) for *BRAF*^V600E^ and *RAS* mutations, as PTC exist as separate BRAF-or RAS-like subtypes^25^. *BRAF*^V600E^ mutations exhibited elevated VAF in PTC relative to ATC, suggesting earlier, preferentially clonal, evolutionary timing.

**Table 1:** Cross-study comparison of common mutations. TP53 and BRAF are among the most frequently mutated genes in ATC and these frequencies show clear trends in relation to degree of differentiation. Numbers indicate mutation frequency (percent of samples) in each cohort; values in brackets indicate median variant allele frequency across all samples in that cohort). ATC: anaplatic thyroid cancer; DTC: differentiated thyroid cancer; PTC: papillary thyroid cancer; HTC: Hurthle cell carcinoma.

|  | Study | ATC | DTC | PTC/HTC |
| --- | --- | --- | --- | --- |
| TP53 | GATCI | 37% | 21% | 1% |
|  | TCGA | N/A | N/A | 0% |
|  | Landa | 70% | 9% | N/A |
|  | Pozdeyev | 48% | 8% | N/A |
|  | Yoo | 44% | 20% | N/A |
|  | Ganly | N/A | N/A | 7% |
|  | Gopal | N/A | N/A | 13% |
| BRAF | GATCI | 21% (11.4%) | 50% (23.6%) | 51% (30.0%) |
|  | TCGA | N/A | N/A | 59% (35.3%) |
|  | Landa | 45% | 36% | N/A |
|  | Pozdeyev | 37% | 59% | N/A |
|  | Yoo | 41% | 27% | N/A |
|  | Ganly | N/A | N/A | 0% |
|  | Gopal | N/A | N/A | 3% |

These patterns of driver mutations were also mirrored for mutational processes. The two prominent mutational signatures activity in PTCs were COSMIC signatures 2 and 13 (AID/APOBEC activity) and 6 (MMR deficiency; **Figure 3F**, **Supplementary Figure 5A**). These were uncorrelated to patient sex or age (Student’s *t*-test, FDR > 0.1), and were detected in ATC. Of the co-occurring DTCs about half showed mutational signatures identical to their ATC counterparts and the remainder showed either higher activity of a specific signature, or a mix of multiple signatures. These data are consistent with a model where ATCs and DTCs evolve from a common precursor, and that specific mutations within that common precursor increase the risk of ATC evolving. This has been proposed previously, based on limited panel-sequencing, but never confirmed with genome-wide systematic subclonal reconstruction. We therefore performed high-depth whole-genome sequencing of paired ATC and co-occurring DTC and used this multi-region data to infer the clonal hierarchy and mutation timing for each patient using a validated subclonal reconstruction pipeline^47,48^ for both SNVs (**Figure 3D**, **Supplementary Figure 6B**; **Supplementary Table 10**) and subclonal CNA status (**Figure 3E**; **Supplementary Table 11**), and validated with high-coverage (>500x) targeted and whole-exome sequencing (**Supplementary Figure 6D**).

As anticipated, each tumor exhibited a distinctive evolutionary history, but every single case shares two features. First, in every single case, co-DTC and co-occurring ATC shared a common clonal origin, with a common ancestor splitting to form separate lineages with significant subclonal diversification (**Figure 3G–P**). This split of the ATC and DTC lineages typically emerged early in tumor evolution: the common ancestor harboured ∼95% of CNAs (**Table 2**), but only 19.1 ± 7.9% of SNVs. Regions of co-occurring DTC were not restricted to a specific CNA subtype, and showed no consistent trends in PGA, total number of CNAs or other characteristics (**Figure 3F**), consistent with a lack of clonal interactions.

**Table 2:** Summary of phylogenetic reconstruction. Multi-sample subclonal reconstruction was performed on nine samples with paired regions of ATC and co-occurring DTC tumor. C = Common; PGA: percent genome altered; CAN: copy number aberrations; ATC: anaplastic thyroid cancer; DTC: differentiated thyroid cancer

|  | SNVs |  |  |  | PGA |  |  |  | CNAs |  |  |  |
| --- | --- | --- | --- | --- | --- | --- | --- | --- | --- | --- | --- | --- |
|  | Trunk | C | ATC | DTC | Trunk | C | ATC | DTC | Trunk | C | ATC | DTC |
| ATCWGS.33 | 3 | 51 | 148 | 104 | 0.00 | 12.35 | 0.06 | 0.00 | 0 | 241 | 4 | 0 |
| ATCWGS.34 | 350 | 26 | 857 | 871 | 0.00 | 32.40 | 0.00 | 0.00 | 0 | 437 | 1 | 0 |
| ATCWGS.35 | 1686 | 0 | 1420 | 1575 | 92.64 | 0.00 | 0.05 | 0.32 | 604 | 0 | 14 | 37 |
| ATCWGS.36 | 364 | 0 | 262 | 2347 | 40.10 | 0.00 | 0.20 | 1.45 | 731 | 0 | 49 | 4 |
| ATCWGS.37 | 517 | 0 | 316 | 1486 | 70.75 | 0.00 | 0.60 | 1.27 | 1207 | 0 | 6 | 3 |
| ATCWGS.38 | 302 | 0 | 852 | 2245 | 13.87 | 0.00 | 71.38 | 0.00 | 150 | 0 | 360 | 0 |
| ATCWGS.39 | 166 | 58 | 376 | 380 | 0.00 | 46.30 | 0.02 | 0.00 | 0 | 1194 | 14 | 1 |
| ATCWGS.40 | 250 | 0 | 1237 | 309 | 22.13 | 0.00 | 0.00 | 0.00 | 238 | 0 | 0 | 0 |
| ATCWGS.41 | 437 | 0 | 979 | 703 | 42.04 | 0.00 | 0.00 | 1.25 | 917 | 0 | 3 | 123 |

In most cases the spatially separated ATC component contained no DTC clones, as in ATCWGS37 (**Figure 3H**) and ATCWGS34 (**Figure 3C**) where in each case the rapidly-growing ATC exhibits only a single subclone, defined as those with a CCF of less than 80%, at our limits of detection. In several cases, we were able to clearly identify a mutagenic field, for example in ATCWGS34 (**Figure 3C**) and ATCWGS33 (**Figure 3J**). In these cases infiltrating stroma harbored common mutations in both ATC and co-occurring DTC, providing clear evidence for a prior mutagenic field from which both tumor components emerge. ATCWGS33 also provides a remarkable example of the complexity of the subclonal interactions between the two tumor components. The tumor diverges from a common field into two branches (subclones 2 & 3, **Figure 3J**). Both the differentiated and anaplastic tumor regions contain clones from both branches of these tumors, but with significant differential evolution: compare subclone 3 (found in DTC at low frequency) to subclone 6 (found in ATC at high frequency).

While several mutations show biases in evolutionary timing (**Supplementary Figure 6B**), with *BRAF* SNVs notably occurring in a clonal or early subclonal population. We compared primary to metastatic ATC for ANPT0021 (**Supplementary Figure 6B**, bottom left panel): the primary ATC exhibits a clonal or high-prevalence subclonal *BRAF* mutation (y-axis) while its cervical metastasis has nearly undetectable levels of this variant suggesting either allelic loss or that the metastasis derived from a low-frequency subclone in the primary tumor. Thus, *BRAF* variants are frequent in both PTCs and co-occurring DTCs, and rare in ATCs. Taken togerhter, these data are consistent with a model where ATCs and DTCs emerge from a common mutagenic field effect. More studies will be needed to validate these findings in tumors from other ATC patients.

## Discussion

The vast majority (>90%) of thyroid cancers are well-differentiated papillary and follicular types with extremely favorable prognoses (>90% survival at 10 years)^49–52^. Yet ATC is rare, and one of the most lethal human malignancies: it is intriguing that the same organ harbors both the most aggressive and some of the most indolent, manageable tumors. The genomic landscape of PTC has been reported to have a low mutational burden^25^, with either single activating oncogene mutations (*BRAF* or *RAS* family genes) or receptor tyrosine kinase gene fusions^25^. The genomic landscape of Hurtle cell carcinoma is similarly quiet, with a small number of chromosome duplications (particularly chromosome 7) and uniparental disomy, and few recurrent point mutations^32,33^. In contrast, the genomic landscape of ATC is complex, with significant inter-patient heterogeneity; individual tumors carry mutations in multiple oncogenes and tumor suppressors, and an overall elevated rate of copy number changes.

ATC frequently occurs in patients with a previous history of DTC or contains co-existing areas of DTC^11–13^, leading to speculation that some ATC evolves *via* dedifferentiation from DTC. However, prior evidence has been limited to analysis of a small number of genes, particularly *BRAF*^44,45^. WXS and CNA profiling of the DTC and ATC components from 21 tumors, combined with WGS profiling of paired ATC and co-occurring DTC components from 9 tumors, conclusively demonstrated that both components share a common genetic origin. High-coverage validation of a subset of cases showed that many ATC driver mutations are subclonal, and exclusive to the ATC component. Multiple somatic SNVs and CNAs are more frequently altered in ATC than DTC, and a subset of these are likely critical drivers of thyroid cancer progression. Functional studies are necessary to determine which mutations lead to the dedifferentiation and increased aggression of well-differentiated thyroid cancer models.

Our subclonal reconstruction analysis suggests that ATCs evolve from a DTC subclone, after accumulation of mostly DNA double-stranded breaks (CNAs). This clone then acquires characteristic additional oncogenic drivers and the majority of its single-stranded breaks (SNVs). The original DTC clone and other subclones in the mutagenic field can continue to exist within ATC as infiltrating stroma, and only the ATC clone harbors metastatic potential. These data suggest a model where both ATC and DTC arise from a non-malignant mutational field effect, where a set of cells accumulate some SNVs and many CNAs through some combination of environmentally-driven and replication-associated mutational processes. Some of these cells then accumulate the characteristic somatic mutations of DTCs and give rise to curable tumors. A different subset accumulates further specific driver mutations, and eventually subclonal driver point-mutations in genes like *TP53*. This model suggests that their evolutionary trajectories may lead to the differential aggressiveness of these related tumor types and the expansive subclonal architecture of ATC. ATC would then emerge from precursors within its co-occurring DTCs by significantly increasing genomic diversity and evolve further as it colonizes new metastatic sites. It is unclear if this model also holds for ATCs not arising in the context of DTCs – WGS of non-malignant thyroid and multi-region ATC specimens will be required to address this question.

Development of the 15 institution Global Anaplastic Thyroid Cancer Initiative (GATCI) was critical to facilitate the characterization of this large number of ATC samples, pathologic review by world experts, and a platform for future collaborative studies. However, there are limitations to this first work, including the depth of sequencing, which may have resulted in lower mutation rates than reported in other studies^17,19,21,23,53^, such as the heterogeneity in platforms used, incomplete clinical data for all patients, and the fact that most samples were collected prior to widespread use of targeted therapy, which may have affected survival outcomes. We are directly addressing these issues in future projects. Despite this, our multi-omic platform study defined the driver landscape and disease subtypes, and identified a long tail of single nucleotide alterations and recurrent copy number variations, and frequent activation of AID/APOBEC mutational process. Furthermore, we delineated the evolutionary relationship between ATC and differentiated thyroid cancers and provided genomic evidence that these two cancer types share a clonal origin. Taken together, our data not only provides a major resource to the thyroid cancer research community, but has led to ongoing work evaluating the functional importance of the novel genes and mutational process that we have identified.

## Methods

### Sample collection and handling

Research Ethics Board approval was obtained at all site locations. Tumors from patients previously diagnosed as ATC were reviewed by at least one study pathologist at each site. Samples meeting the American Thyroid Association diagnostic criteria for anaplastic thyroid cancer^9^ were selected for study including both fresh frozen and formalin fixed paraffin embedded (FFPE) samples (**Supplementary Table 1**). All WXS tumors were from fresh frozen samples or FFPE while matched normal subjected to WXS were from FFPE. All tumor samples subjected to WGS were from FFPE while and matched normal were from blood tissue. Well-differentiated and/or poorly differentiated components were macro-dissected when present separately from the undifferentiated ATC components using 1 mm punches. Fresh tumor was prospectively collected at four institutions with matched blood collected in the majority of cases. DNA was extracted using Qiagen kits. Where available, samples were subject to centralized pathology review to estimate tumor purity and confirm diagnoses. PTC samples (n = 115 from 112 patients, **Supplementary Table 1**) were selected from the tissue bank at MD Anderson Cancer Center based on age and tumor status (organ confined, regional involvement, or regional and distant metastasis). DNA was isolated and processed as described previously^54,55^.

### Detection of Copy Number Aberrations

Affymetrix OncoScan FFPE assays were used to evaluate a total of 157 samples (including 99 ATCs, 24 samples from co-occurring well (20) or poorly (4) differentiated regions, 1 co-occurring cervical ATC metastasis, 20 normal thyroid samples and 13 ATC-derived cell lines) for somatic copy number aberrations (CNAs). Analysis was performed using .OSCHP files generated by OncoScan Console 1.1 using either build 33 of the NetAffx annotation (FFPE samples) or a custom reference consisting of 119 normal blood samples from male patients with prostate cancer, 2 normal blood samples from females with anaplastic thyroid cancer and 10 female HapMap cell line samples (fresh frozen samples and cell lines). BioDiscovery’s Nexus Express Software was used to call copy number aberrations using the SNP-FASST2 algorithm with default parameters, with manual re-centering performed as required. Nine samples (six normal tissues, one ATC and two well-differentiated tumor components) were removed due to poor quality and the remaining data evaluated for recurrence. Cellularity and ploidy of tumor samples was assessed using ASCAT (v2.5) package in R. LRR and BAF values were obtained from the .OSCHP files. Tumor ploidy and aberrant cell fraction estimates for each sample were obtained using either predicted germline SNPs or by leveraging matched normal arrays where available.

PTC samples (113 tumors), along with an additional 13 ATCs, were processed using Illumina Human Omni2.5-8 or OmniExpress12v1-1 beadchips, as described previously^54,55^. Briefly, DNA was denatured, amplified, enzymatically fragmented and hybridized for 16-24 hours at 48°C. Beadchips were imaged using the Illumina iScan system. For improved CNV analysis, B allele frequencies (BAF) were calculated and log2 R ratios (LRR) were extracted after re-clustering the raw data using GenomeStudio cluster algorithms. BAF and LRR were then loaded into Nexus to identify somatic DNA copy number aberrations. UCSC’s command-line liftover tool (v359) was used to transfer resulting CNAs to GRCh38 coordinates.

CNAs from both platforms were combined, and UCSC’s command-line liftOver tool (v359) was used to convert segment positions to hg38 coordinates (using the hg19-to-hg38 chain file downloaded from UCSC). Gene level copy number aberrations for each patient were identified by overlapping CN segments with Ensembl annotation (Ensembl release 84). Percentage of genome altered (PGA) was calculated for each sample by dividing the number of base-pairs involved in a copy number change by the total length of the genome. GISTIC2.0 (v2.0.22)^56^ was used to study the recurrence of gene level CNAs. For each sample, a profile was created that segmented each chromosome into regions with neutral, CN loss, and CN gain events. The average copy number intensity for each segment was obtained from the SNP array analysis. Chromosome-level events were defined as a gain or loss of 25% of the chromosome for each sample.

Subtype discovery was performed on CNA calls for 110 ATCs using consensus clustering to determine the optimal number of clusters and class membership for each sample by stability evidence. Specifically, the ConsensusClusterPlus.custom (v1.8.1) package for R was used to evaluate a wide variety of clustering methods for distributing samples into k clusters (where k is every value from 2 to 10) using 1000 sub-samplings of 80% of the cohort. The method which produced the most stable groups used hierarchical clustering using a Euclidean distance similarity metric with modified Ward’s minimum variance method (ward.D) and 5 clusters.

Data visualization was performed in the R statistical environment (≥ v3.2.3). Venn diagrams were created using the VennDiagram package (v1.6.17) and all other data visualizations were generated using the BPG (v5.8.8), lattice (v0.20-35) and latticeExtra (v0.6-28) packages.

### DNA Whole-Exome Sequencing (WXS)

Tumor samples (and matched normal tissue where available) from 139 patients (including 139 ATCs with 20 co-occurring regions of well or poorly differentiated tissue, 1 cervical ATC metastasis) and 13 ATC-derived cell lines (total = 173 tumors) obtained from eight centres were sequenced at one of six sequencing facilities (**Supplementary Table 1**). Raw FASTQ files were provided for a subset of samples while the remainder were provided in BAM format. In the latter case, read extraction was performed using the SamToFastq component of Picard tools (v1.121) to produce FASTQ files. Data were aligned to the GRCh38 human reference genome, including available ALT and decoy alleles using BWA-mem (v0.7.15)^58^. The reference files were downloaded from NCBI (GCA_000001405.15; release date 2013/12/17). Duplicate reads were marked using Picard tools (v1.121). As the exome capture kit regions were provided in hg19 coordinates, USCS’s web-based liftover tool (https://genome.ucsc.edu/cgi-bin/hgliftover) was used to convert regions to GRCh38 positions. The Genome Analysis Toolkit (GATK v3.5.0)^59^ was used for local realignment and base quality recalibration, with regions limited to those targeted by the applicable exome capture kit (**Supplementary Table 1**). As hg38 has not yet been widely adopted, a beta-version of the hg38bundle containing known variants was downloaded from the Broad (ftp://ftp.broadinstitute.org/bundle/hg38/hg38bundle/).

GATK’s HaplotypeCaller (v3.5.0) was used to identify germline SNPs and indels using default parameters, however with --output_mode EMIT_VARIANTS_ONLY --emitRefConfidence GVCF --standard_min_confidence_threshold_for_calling 50. GenotypeGVCFs and SelectVariants tools were used to format variant calls, followed by VariantFiltration with the following criteria: (QD < 10.0 || FS > 60.0 || MQ < 40.0 || DP < 50 || SOR > 4.0 || ReadPosRankSum < -8.0 || MQRankSum < -12.5), (MQ0 >= 4 && ((MQ0 / (1.0 * DP)) > 0.1). Variants were annotated using SnpEff (v4.3)^60^ and filtered to keep any position with coverage in the 1000 genome project that had a population allele frequency <1%. These were then filtered to keep variants in clinVar and to remove synonymous variants. Finally, for patients with multiple tumor components, germline variants present in a single component were labelled false positives and removed. Remaining variants were searched against a list of relevant cancer predisposition genes^29^ and checked for recurrence.

Coverage was estimated across target regions using BEDTools (v2.18.2) (**Supplementary Figure 2a**). For tumor/normal pairs, ContEst (v1.0.24530)^61^ was used to estimate cross-sample contamination, using the above generated germline SNPs. As the necessary population allele frequencies were not yet available using GRCh38 coordinates, allele frequencies were obtained from HapMap (hg19) and added to the GRCh38 HapMap file, with variants matched across builds using rsID. Seven samples demonstrated contamination >3% and were removed from downstream analysis (**Supplementary Figure 2b**).

For samples with matched normal tissue, somatic SNVs were predicted using SomaticSniper (v1.0.5.0)^62^, as described previously^6^. A panel of normals (PoN) consisting of the 83 normal thyroid samples was generated using MuTect’s (v1.1.7) artifact detection mode; variants detected in two or more normal samples were included in the PoN. For unmatched samples, SNVs were identified using MuTect (v1.1.7)^63^ with PoN, with dbSNP (build 150) and COSMIC (v74) filters; COMSIC variants present in five or more samples within the PON were discarded. To combine samples processed at different sequencing facilities, predicted somatic variants were filtered to regions appearing in all exome capture kits used (**Supplementary Table 1**). Overlapping regions across kits were identified using bedtools:multiIntersectBed (v2.24.0), resulting in coverage of roughly 26 Mbp. Recurrence analysis was performed using RecSNV (v2.1.5). Briefly, all variants were annotated using Annovar (v2016Feb01) with comprehensive filtering performed to remove known germline variants using previously described datasets^6^; any variant present in COSMIC (v82) was retained, regardless of presence in other datasets. Significance of recurrent non-synonymous mutations was assessed using SeqSig (v3.7.8)^6^, using a required mutation threshold of 6 samples across the cohort per gene, individual sample weights of -log_10_(x) + log_10_(1-x) [where x is the background mutation rate per sample], with p-values calculated using the exact distribution, followed by FDR correction for multiple testing. Genes containing variants in multiple samples were carried forward. Genes harbouring recurrent SNVs were visualized using lollipop plots (**Supplementary Figure 3**). For each gene, protein domain information was obtained from NCBI and pfam (2018-02-07), showing the most well conserved domains where feasible.

PTC samples (115 tumors with matched normal and 1 metastatic lymph node) were processed separately, as described previously^54,55^. Briefly, DNA was prepared and processed for WXS at the Human Genome Sequencing Center (HGSC) in Baylor College of Medicine (BCM). Reads were aligned to GRCh37 by BWA^58^, duplicate reads were marked by Picard tools and BAMs realigned/recalibrated by GATK^59^. Somatic SNVs were identified using Atlas-SNP; filtering was applied to ensure variants had a minimum of 4 high-quality support reads and a minimum VAF of 0.08. For comparison to the above ATC cohort, the resulting SNVs were transferred to GRCh38 coordinates using UCSC’s command-line liftOver tool (v359).

### Deep Targeted IonTorrent Sequencing

A total of 1,140 variants were selected for validation based on recurrence within the initial dataset of 30 ATCs. Primer design generated an AmpliSeq-custom panel for 1125 of these variants (995 target regions). A subset of samples was used for targeted validation by IonTorrent sequencing including 30 ATCs (13 with matched normal, 3 with matched co-occurring DTCs and 1 cervical ATC metastatic tumor), 13 ATC-derived cell lines and an additional 7 ATCs without WXS data. DNA for each sample was shipped to GeneDX for sequencing. Raw FASTQ files were provided and reads were aligned to GRCh38, as described above however without marking of duplicate reads. Targeted sequences were converted from hg19 to GRCh38 coordinates using liftOver (UCSC). Read lengths were extracted from FASTQ files and visualized against target region length (**Supplementary Figure 2c**). Coverage was estimated across target regions using BEDTools^64^ (v2.18.2) (**Supplementary Figure 2d**).

Base counts at target positions were assessed and used to calculate variant allele frequencies (**Supplementary Figure 2e**). Validation was performed as described previously^65^. Briefly, variant positions were used, along with either the GATK-processed WXS BAMs or the IonTorrent BAMs to generate a modified pileup file indicating the base counts at each SNV position. For T/N pairs, metrics including a χ^2^ test of the base-count distribution between tumor and normal at each position followed by Bonferroni adjustment of the p-values (padj < 0.25; **Supplementary Figure 2f left panel**), Euclidean distance between variant allele frequencies (> 0.15; **Supplementary Figure 2f right panel**), z-score of the sample coverage across target positions (< 2 standard deviations from the mean for tumor and normal separately), and ternary allele proportion for normal samples to quantify potential false positive variants (< 0.05) were used to classify variants as somatic or non-somatic. For tumor only samples, similar metrics were used, however comparing Deep IonTorrent and WXS values to classify read-counts as similar or different between the platforms. Accuracy, sensitivity and specificity were assessed for each sample (**Supplementary Figure 2g**). Variants identified as false-positives using this method were excluded from WXS analyses.

### DNA Whole-Genome Sequencing (WGS)

Whole-genome sequencing (WGS) was performed on nine ATCs, each with paired co-occurring DTCs and matched normal. Genomic DNA was isolated from FFPE samples and sequenced at The Center for Applied Genomics (Hospital for Sick Children, Toronto, ON). FASTQ files were downloaded and processed using the methods described above. Somatic SNVs were identified using SomaticSniper (v1.0.5.0)^62^, as described above and SCNAs were identified using MutationSeq (v4.3.7) and TITAN (v1.20.1)^66^, followed by multi-sample subclonal reconstruction using PhyloWGS (v1.5)^67^. Clonal mutations are defined as mutations that are present in all tumour cells (CCF >= 0.8) of a tumour sample or biopsy. A subclones is defined as those that is a descendent of the most recent common ancestor of the tumour sample and with CCF < 0.8 in at least one tumour region.

### Validation of TERT promoter mutations by PCR

To determine the mutational status of the TERT promoter, we used a nested PCR strategy with our FFPE patient samples. Genomic DNA in this region is 78% GC rich and it was necessary to use PCR additives to amplify the region. The primers were designed and tested *in silico* using PRIMER3 software. The first round PCR was performed with primers hTERTF x hTERTR under the following conditions: initial denaturation at 98°C for 30 seconds followed by 36 rounds of 98°C denaturation for 10 s and combined annealing and extension at 72°C for 25 sec. We employed the Q5 high fidelity DNA polymerase (New England Biolabs; NEB) and the supplied 5x PCR buffer and GC enhancer (NEB) to amplify a 473 bp region. The second round of PCR was initiated with 0.2 ul template taken directly from the first round using primers TERT5 F x TERT5 R again with the addition of the GC enhancer and the same conditions increased to 40 cycles. PCR reactions were run on a 2% TBE agarose gel and the expected 192 bp product was excised and gel purified using a Monarch Gel Extraction Kit (NEB) following the product protocol. The purified fragments were Sanger sequenced at the London Regional Genomics Facility with betaine (1M) added to the reaction. Chromatograms were aligned and compared against the TERT sequence.

### RNA Sequencing

Total RNA was isolated from 13 ATC cell lines and 8 tumor samples using Qiagen AllPrep DNA/RNA kits and shipped to The Center for Applied Genomics (Hospital for Sick Children, Toronto, ON) for sample processing using random primers and sequenced using the manufacturer’s protocols. Specifically, sequences were generated using paired-end, strand-specific methods. Data were provided in BAM format; FASTQ files were re-generated using picard (v1.141) CleanSam and samtools (v1.4) fastq functions. A second set of 16 patient tumors were processed as described previously^55^. Briefly, frozen tissue was thawed in RNAlater™-ICE Frozen Tissue Transition Solution (ThermoFisher Scientific) to enable easy extraction of high-quality RNA. Total RNA was prepared using TRIzol reagent (Life Technologies) according to the manufacturer’s instructions. Poly-A+ Illumina RNA-Seq libraries were prepared and paired-end sequencing performed using the Illumina HiSeq 2000 and HiSeq 2500 platforms at the HGSC (Baylor College of Medicine, Houston, TX).

For mRNA abundance, top, level DNA sequences and gene annotations (GTF) were downloaded from ensemble.org (release 88) and used to generate reference files needed for RSEM (v1.3.0)^68^, using the STAR (v2.3.3a)^69^ alignment tool; transcripts per million (TPM) values are available in **Supplementary Table 2**. Fusion detection was performed using fusioncatcher (v0.99.7c)^38^. Detected fusions are shown in **Supplementary Table 5**; these were compared against the COSMIC fusion database (2019-12-13).

### Statistical Analysis

Non-negative matrix factorization (NMF) was used to assess the contribution of various trinucleotide signatures within our ATC and PTC cohorts separately. NMF was performed in R (v3.4.1) using the NMF package (v0.20.6) using counts of abnormal trinucleotides from undifferentiated tumors. Rank estimation metrics, combined with visual inspection of consensus clustering maps, suggested the presence of five trinucleotide signatures for ATC, and two for PTC. Signatures were mapped to previously described trinucleotide signatures from COSMIC (downloaded from COSMIC on 2017-12-08) using the consensus of Euclidean distance, Pearson’s or Spearman’s correlation, followed by the Hungarian method for linear sum assignment^70^ using the clue (v0.3-54) package for R. Spearman’s correlation did not identify any associations between these 5 signatures and clinical covariates or CNA clusters.

Gene-wise mutation frequencies from the current dataset were compared to previous studies of thyroid cancer, including a set of 22 ATCs by WXS published by Kunstman *et al*.^15^, 84 PDTCs and 33 ATCs by targeted-sequencing published by Landa *et al*.^17^, 182 PDTCs (including PTC, FTC and HTC) and 134 ATCs by targeted-sequencing published by Pozdeyev *et al*.^21^, 27 ATCs and 15 PDTCs from Yoo *et al*.^23^, 2 independent studies of 32 and 56 HTCs respectively by WXS published by Gopal *et al*. and Ganly *et al*. and 496 PTCs from TCGA (MAF downloaded from GDC on 2017-11-02)^25^ (**Supplementary Table 9**). Where sufficient annotation was available, samples from each study were filtered to remove metastatic or other non-primary thyroid tumors. Variants from the TCGA PTC cohort were filtered to keep only those with predicted functional relevance with an additional coverage threshold applied to the MAF (minimum 10 reads in both tumor and matched normal). SNV data were collapsed to gene-level and proportion tests used to evaluate differences between studies (ATC or co-occurring DTC (GATCI) *vs*. TCGA PTC), followed by FDR adjustment for multiple testing.

Similar comparisons were made for CNAs, again using TCGA PTC data (masked copy number segment data downloaded from GDC on 2018-01-29). TCGA data were converted to ternary CN values and annotated with overlapping genes to produce a gene by sample matrix (primary tumor samples only). Proportion tests were again used to assess differences in population frequencies for each gains/losses separately, with FDR-adjustment (**Supplementary Table 11**).

Well or poorly-differentiated co-occurring tumor components were obtained from 24 patients and a single patient provided both an ATC and metastatic tumor obtained from the cervical region. For CNA analysis, data were available for 20 co-occurring DTC tumor components as well as ATC. Hypergeometric tests were used to evaluate the CNA overlap across regions within each patient [P(actual overlap ≥ expected overlap)] (**Supplementary Table 11**). Similar analyses were performed for SNV overlap (16 patients with multiple components; **Supplementary Table 10**).

Clinical variables were assessed for associations with each other using χ^2^ independence tests to assess co-linearity, and with overall survival to identify which should be used as covariates in downstream models. Age, nodal metastases and treatment methods (surgery, radiotherapy and chemotherapy) were all individually associated with survival (FDR < 0.1). Treatment methods showed considerable overlap, as did nodal and distant metastases (FDR < 0.1). Therefore covariates including age, nodal metastases and surgery were used for survival analyses. Overall survival was evaluated using either CNA or SNV features. For copy number status, of 961 genes identified by GISTIC, 723 were CN altered in 10% of the cohort with available survival data (n = 83) and were collapsed to 187 genomic regions. Regions were collapsed to binary status where reasonable, such that the CN type with 4 or fewer cases was merged with neutral. Further, CN status of known driver genes, with a minimum recurrence of 10 samples, were also tested (n = 108). Associations with survival were evaluated using either log-rank (for ternary CN status) or Cox proportional hazards regression model with linear adjustments for covariates described above (binary CN status). Binary cases which failed the assumptions for the Cox model were repeated using log-rank tests. FDR-adjustment of the p-values was applied for multiple testing correction (**Supplementary Table 7**). For point mutations, 101 patients had available survival data. Feature reduction was performed to remove low frequency variants (≥ 5 patients); this reduced the feature set from 14,543 genes to 514 recurrently altered genes. Associations with survival were using a Cox proportional hazards regression model with linear adjustments using covariates described above, or log-rank test where the Cox model failed its assumptions. FDR-adjustment of the p-values was applied for multiple testing correction (**Supplementary Table 6**).

In addition to gene-wise features, clinical and survival associations were also evaluated using PGA or SNVs/Mbp. Associations with clinical variables were performed using Wilcoxon rank sum tests. For survival analyses, PGA and SNVs/Mbp were median dichotomized and a Cox proportional hazards regression model with linear adjustments using covariates described above was performed. For SNV mutational density, only data from samples with a matched normal were used.

Finally, mutational features were combined and examined for overlaps. Specifically, pairwise comparisons were made between all combinations of CNA features (key GISTIC genes, numbers of CNAs/gains/losses, average CNA length, PGA and CNA subtypes) and SNV features (top recurrently mutated genes, SNVs/Mbp and trinucleotide signatures using either a χ^2^ test, Spearman’s correlation or non-parametric Wilcox or Kruskal-Wallis rank sum test with FDR adjustment of the p-values. Similar comparisons were made between RNA abundance (TPM) and CNA/SNV/fusion status using Wilcox rank sum tests.

### Data availability

All ATC WXS, WGS and SNP array data is available in EGA under accession EGAS00001002234.

## Data Availability

All data will be available via request to ACN

## Acknowledgements

The authors thank all members of the Boutros and Nichols’ laboratories for helpful suggestions and technical support.

## Competing financial interests

All authors declare that they have no conflicts of interest.

## Materials & Correspondence

Please contact Dr. Paul Boutros or Dr. Anthony Nichols with any correspondence or material requests.

## Funding Sources

This work was supported by Canadian Institutes of Health Research grant MOP 377832 to ACN and PCB, MOP 145586 to PCB and by the Government of Canada through Genome Canada and the Ontario Genomics Institute (OGI-125). NA, BN and DB were supported by a Julius Edlow Pilot Research grant. PCB was supported by CIHR and Terry Fox Research Institute New Investigator Awards and by the Ontario Institute for Cancer Research to PCB through funding provided by the Government of Ontario. This work was further supported by National Institutes of Health and Medical Research Grants R01 CA136665 (to JAC and RCS), Florida Department of Health Bankhead-Coley Cancer Research Program Grant FL09B202 (JAC and RCS), Mayo Comprehensive Cancer Center Grant P30CA01508343 (RCS), UCLA Comprehensive Cancer Center Grant P30CA016042 (PCB), by the NIH/NCI through support from the ITCR (1U24CA248265-01 (PCB), as well as support from Alfred D. and Audrey M. Petersen (RCS), the Francis and Miranda Childress Foundation Fund for Cancer Research (JAC), the John A. and Bette B. Klacsmann Fund for Cancer Research at the Mayo Clinic in Florida (JAC), the Bruno V. and Bruce E. Zanoni Endowed Research Fund (JAC), the Betty G. Castigliano Fund in Cancer Research Honoring S. Gordon Castigliano, MD, Cancer Research at the Mayo Clinic Florida (JAC), and the Dewitt C. (Dash) Goff Fund for thyroid cancer research (JAC). This work was also supported through generous donations by: 1) the Marty Schaffel Thyroid Cancer Research Fund (MD Anderson Cancer Center, Houston, Texas), 2) London Health Sciences Foundation (Robert and Sheila Wilkes, Betty and Harry Ostrander, Mervyn and Joyce Dietz) and 3) the Woodstock Foundation (Laila, Arnold, Mario, Andrea, Tony and Tomassina Spina, Susan and Bill George, Piero and Maria Manzini, Henry and Rina Deroo, Harry and Shani Loewith, Franco and Carol Castellucci, Tony and Bill Van Haeren, Billy and Jennifer Stevanovich, Tom and Pat Baird, Christine and Jeff Nichols, Cliff and Linda Zaluski, Thomas Vandertuin, Laurie and Paul Green, Kelli and Mike Koopman). ACN was supported by the Wolfe Surgical Research Professorship in the Biology of Head and Neck Cancers Fund.

## Supplementary Figures

**Supplementary Figure 1:**
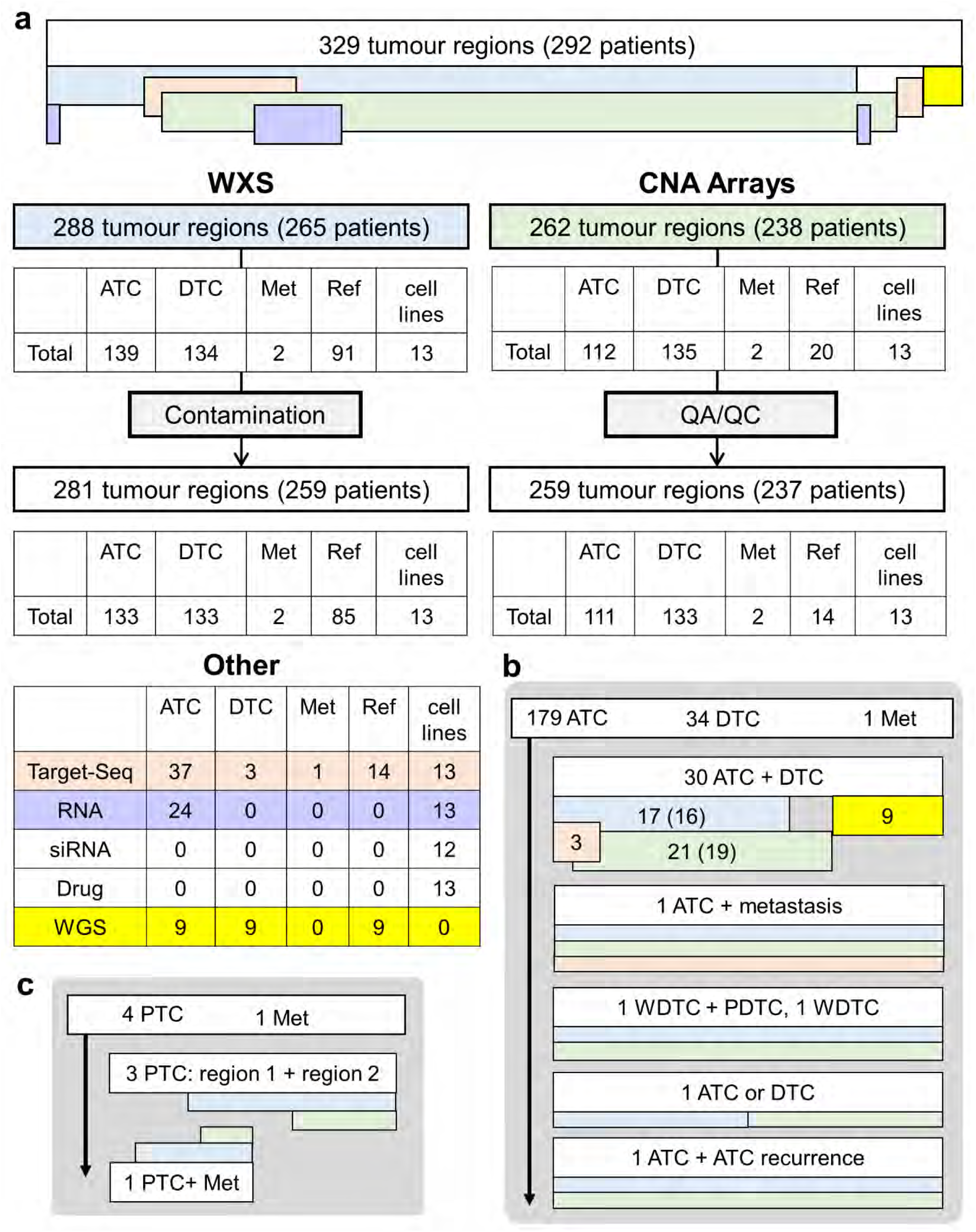
Sample summary. **A)** Breakdown of samples processed by copy number aberrations (CNA) arrays, whole exome sequencing (WXS), deep targeted IonTorrent sequencing, RNA sequencing, WGS, before and after removal of poor quality or contaminated samples. A subset of tumors provided both an undifferentiated (ATC) and co-occurring differentiated component, metastatic (Met) tumor or matched reference sample. **B)** Breakdown of tumors with multiple components: of the 34 co-occurring DTC components available, 30 were processed alongside a paired ATC region (numbers in brackets indicate remaining pairs after QC); one case of both a well-differentiated and poorly differentiated tumor component (no ATC); one case of co-occurring DTC alone; one case of ATC processed by WXS only, with paired co-occurring DTC processed by OncoScan only. **C)** Four patients with PTC had multi-regional interrogation: in three cases, two regions of the primary, and in the fourth the primary tumor and a lymph node metastasis were analyzed.

**Supplementary Figure 2:**
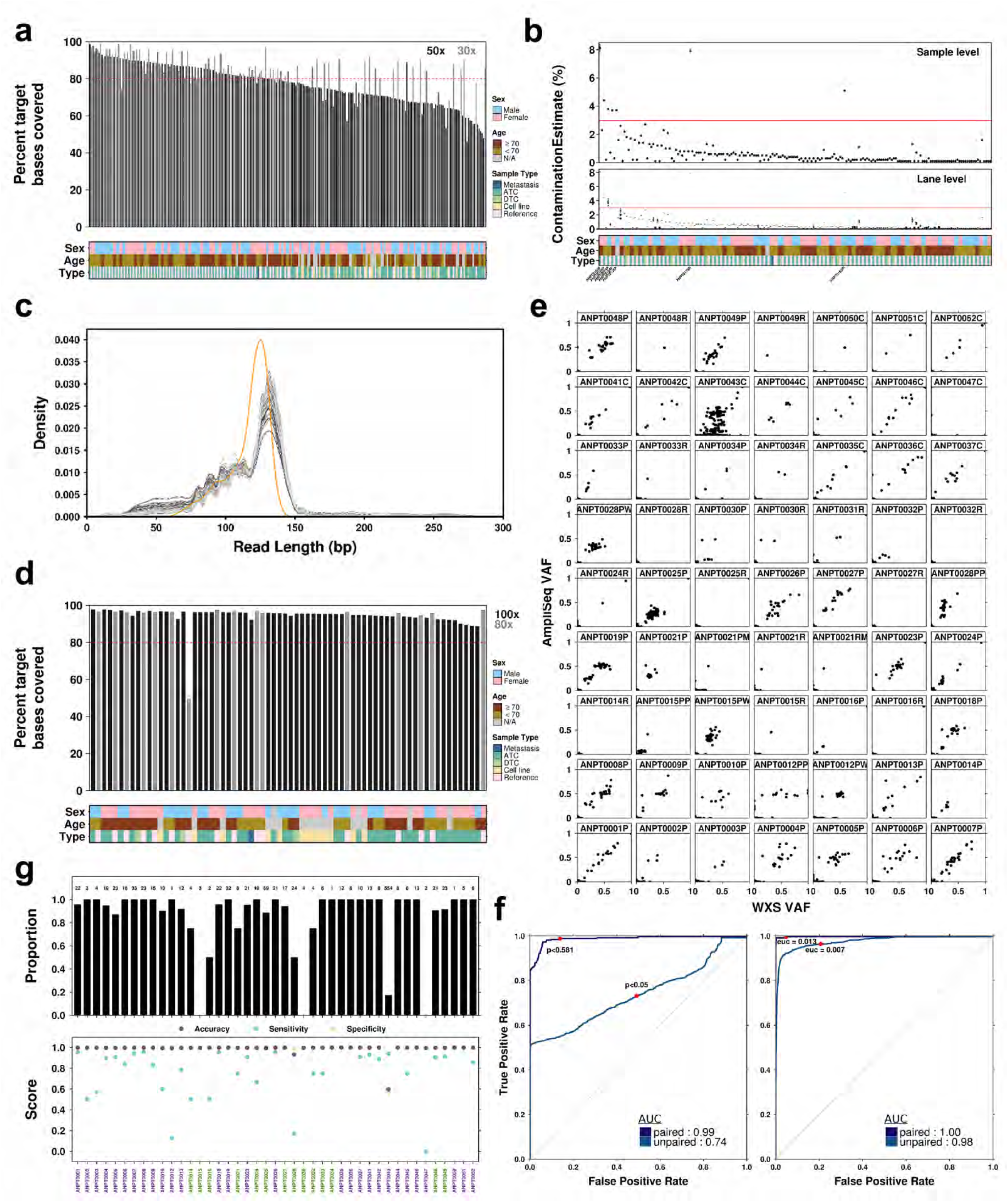
WXS quality metrics and IonTorrent validation. **A)** Percent of target bases from the WXS used that demonstrated 50x (tumor) or 30x (normal) coverage. **B)** ContEst was used to assess cross-sample contamination (T/N pairs only); both sample-level (top) and lane-level (bottom) estimates were obtained where possible. Samples with a contamination estimate >3% were excluded from downstream analyses (n = 7 samples removed). In both cases, samples are ordered from highest to lowest (based on the ATC component), with additional components to the immediate right of these for each patient. For example, in **B)** ANPT0165P has low contamination such that it near the middle, while a poorly-differentiated component from the same patient (ANPT0165PI, immediately to the right of ANPT0165P) demonstrated high contamination by ContEst and was removed from downstream analyses. **C)** Distribution of read length (grey) for each sample or target region length (orange). **D)** Percent of target bases from the AmpliSeq-custom capture design used for IonTorrent sequencing that demonstrated 100x (tumor) or 80x (normal) coverage. **E)** Comparison of variant allele frequencies (VAF) obtained from WXS and IonTorrent targeted sequencing validation for each sample. **F)** AUC for classification of each variant position as somatic/non-somatic using p-value from a Pearson’s χ^2^ test comparing the base counts between tumor and normal or IonTorrent targeted sequencing and WXS (tumor only) (left) or Euclidean distance (right). **G)** Validation metrics: (top) proportion of variants called as somatic in WXS that were successfully validated (total numbers appear above each bar); (bottom) accuracy, sensitivity and specificity for each sample; x-axis label colours indicate T/N (green) or T only (purple).

**Supplementary Figure 3:**
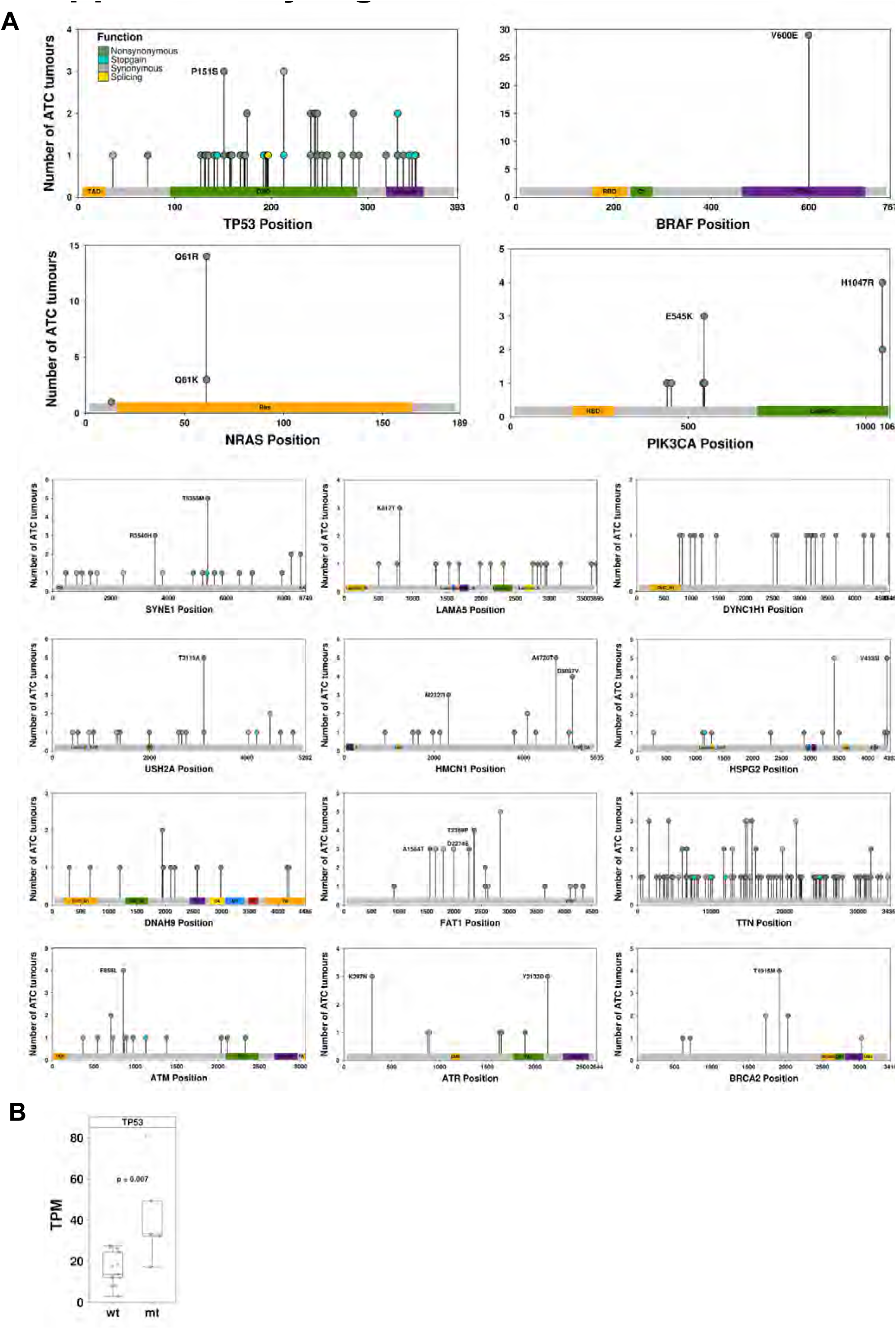
SNV Distribution in recurrently mutated genes. **A**) The position and recurrence of point mutations across the ATC cohort within recurrently mutated genes. Individual non-synonymous variants found in 3 or more patient tumors are labelled with the predicted amino acid change. **B)** Non-synonymous variants in *TP53* were associated with significantly elevated mRNA abundance by RNA-Seq (Wilcoxon rank sum test, p = 0.0067).

**Supplementary Figure 4:**
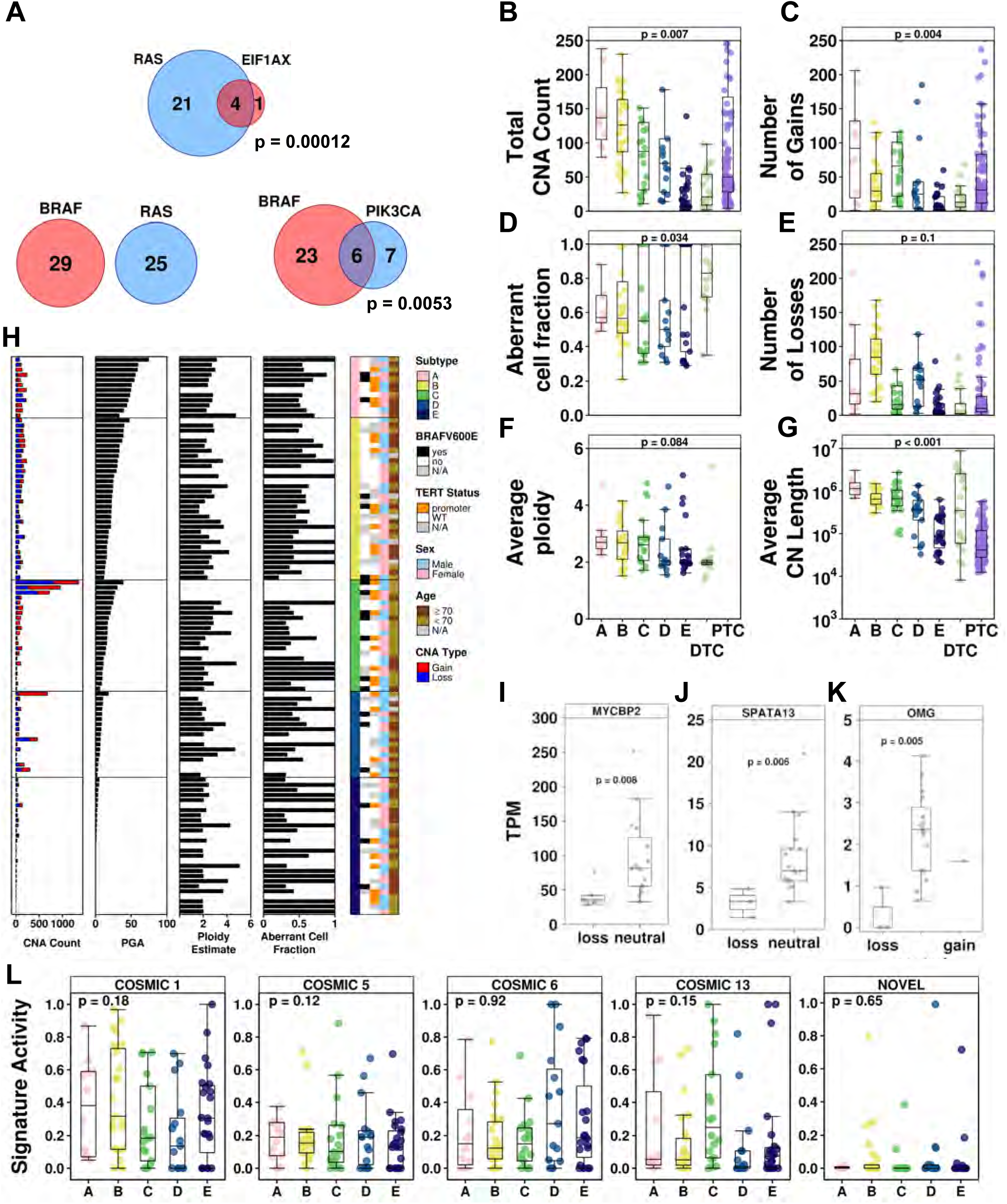
Landscape of Somatic Mutations in ATC. **A)** Point mutations in *EIF1AX* tend to co-occur with *RAS* mutations, as do those in *BRAF* and *PIK3CA*. By contrast, somatic SNVs in BRAF and RAS genes were mutually exclusive. Distribution of **B)** total number of copy number aberrations (CNAs), **C)** number of gains, **D)** aberrant cell fraction (purity), **E)** number of losses, **F)** estimated ploidy, or **G)** average CNA length for patients within each subtype; p-values from one-way ANOVAs. **H)** ATC samples evaluated on CNA arrays to generate a number of metrics for each patient, including CNA count (gains/losses), percent genome altered (PGA), ploidy and purity estimates. Samples are ordered by PGA within each subtype. RNA-sequencing of primary ATC tumors was used to evaluate mRNA abundance of genes identified by GISTIC as being significantly affected by copy number deletions. Wilcoxon rank sum tests were used to contrast mRNA abundance (TPM) between tumors with a CN deletion to those with neutral/gain, followed by FDR adjustment of the p-values. Three genes were found to have significantly different mRNA abundance between groups: **I)** *MYCBP2* (in Figure 1J part of GISTIC segment labeled *KLF5*), **J)** *SPATA13* (part of the *BRCA2* segment) and **K)** *OMG* (part of GISTIC segment labeled *NF1*). **L)** One-way ANOVAs were used to assess the relationship between each trinucleotide signature and CNA subtypes; no associations were detected.

**Supplementary Figure 5:**
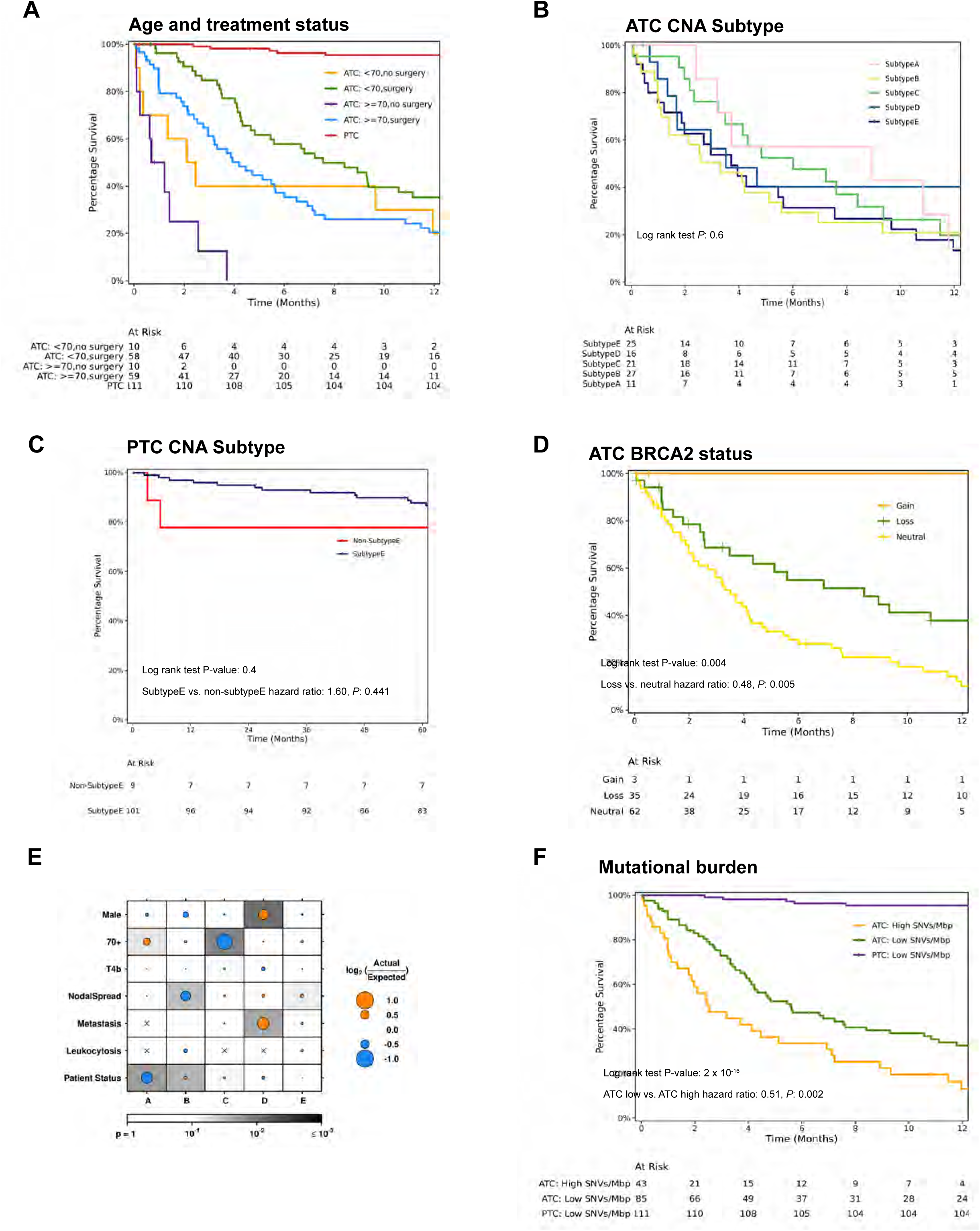
Additional survival analysis. **A)** Age and surgery were associated with overall survival. **B)** ATC CNA subtypes were not associated overall patient survival, howvever copy number deletions of *BRCA2* were associated with superior patient survival (log-rank and Cox proportional hazards tests, **D**). **C)** Overall survival of PTC cases by CNA subtypes **E)** Enrichment of patients with various clinical variables within each ATC-associated can subtype; dot size indicates fold-change (log_2_[overlap_actual_ / overlap_expected_]) within a cluster (x-axis labels) with the specified variable classification (y-axis labels), with orange indicating over-enrichment and blue indicating under-enrichment, while background shading indicates p-value from the hypergeometric test (looking at over-or under-enrichment as defined by dot colour), after accounting for missing data. For example, Subtype E includes patients with increased age and tumor aggressiveness (higher rate of T4b relative to T4a disease, nodal metastases, distant metastasis and leukocytosis). Subtype C has younger patients with fewer T4b than expected by chance alone. Pearson’s χ^2^ test did not identify any difference in distribution of any of these variables across subtypes. **F)** Low ATC SNV mutational density (SNVs/Mbp) was associated with better survival (Cox proportional hazards test); SNVs/Mbp defined as greater than 10SNVs/Mbp. PTC tumors all had low mutational density and experienced superior survival to ATC patients.

**Supplementary Figure 6:**
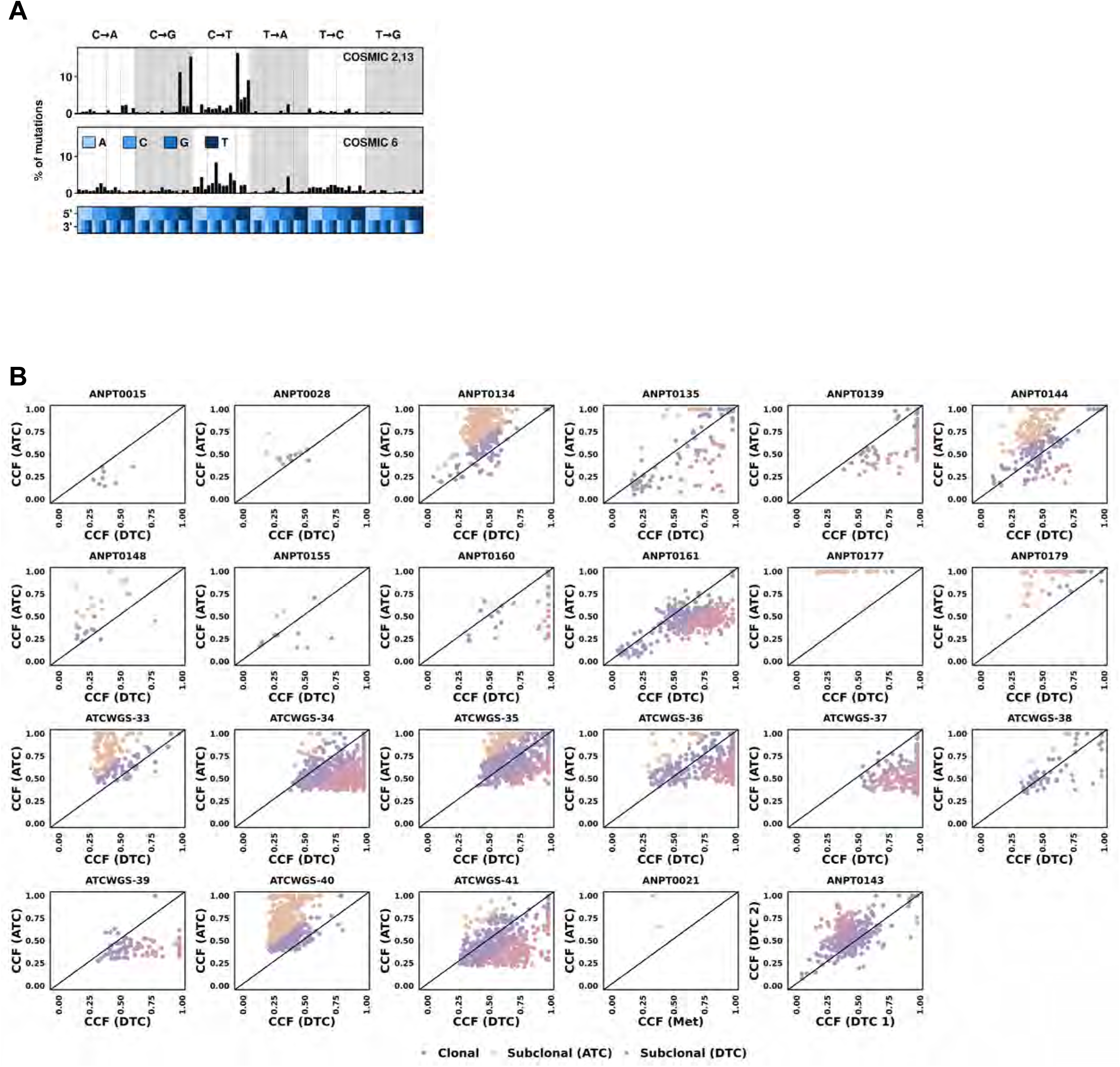
Mutational Timing and Evolution of ATC. **A)** NMF identified two trinucleotide signatures within 112 papillary thyroid cancers (PTCs). Within each signature, the percent of mutations within the cohort presenting each base change (broken ddown by trinucleotide context). **B)** Variants with ≥10x coverage were selected in patients with WGS for both ATC and paired co-occurring differentiated thyroid cancer (DTC).Cancer cell fraction [CCF] were compared to assess tumor evolution. Purple indicates clonal variants (similar CCF), while pink and orange represent co-occuring DTC and ATC subclonal regions respectively. Black points indicate known driver genes.

## Supplementary Tables

**Supplementary Table 1: Sample Characteristics**

a) Clinical characteristics and summary findings of thyroid tumors and cell lines. b) Breakdown of paired ATC and co-occurring differentiated thyroid cancer (DTC) components.

**Supplementary Table 2: RNA abundance in ATC tumors and cell lines**

RNA-sequencing was performed on 24 primary ATC tumors and 13 ATC cell lines; data were normalized and expression values (transcripts per million, TPM) calculated using RSEM and STAR.

**Supplementary Table 3: Germline SNVs in cancer predisposition genes**

Germline SNVs were identified in ATC tumors with match normal only. Results were filtered to remove common variants (<1% in 1000G); SNVs identified in known driver genes are shown.

**Supplementary Table 4: CNA subtype associations with clinicalgenomic characteristics**

Copy number aberrations (CNA) subtypes in ATC were evaluated for associations with patient clinicogenomic characteristics.

**Supplementary Table 5: RNA fusions in ATC**

**a)** RNA-sequencing was performed on 24 primary ATC tumors and 13 ATC cell lines. Fusioncatcher was used to identify gene fusions; results were filtered to remove fusions with low evidence (required at least two reads). b) Fusions involving known driver genes with corresponding RNA abundance (TPM) for affected genes.

**Supplementary Table 6: SNV associations with overall survival in ATC**

Genes identified as recurrently altered in ATC were evaluated for associations with overall patient survival using time-to-event modeling.

**Supplementary Table 7: CNA associations with overall survival in ATC**

Copy number aberrations (CNA) segments identified by GISTIC as being significant in ATC were evaluated for associations with overall patient survival using time-to-event modeling.

**Supplementary Table 8: Cross-study analysis of gene-wise CNA frequencies**

Gene-wise copy number aberrations (CNA) frequencies were calculated for 111 ATC, 24 co-occurring differentiated thyroid cancer (DTC) and 112 papillary thyroid cancer (PTC) primary tumors. These were contrasted to mutation frequencies obtained from TCGA (PTC, n = 505).

**Supplementary Table 9: Cross-study analysis of gene-wise SNV frequencies**

Gene-wise SNV frequencies were calculated for 132 ATC, 19 co-occurring differentiated thyroid cancer (DTC) and 112 papillary thyroid cancer (PTC) primary tumors. These were contrasted to mutation frequencies obtained from TCGA (PTC, n = 481), Kunstman *et al*. (ATC, n = 22), Landa *et al*. (ATC, n = 33; pooly differentiated thyroid cancer [PDTC], n = 78), Pozdeyev *et al*. (ATC, n = 134; PDTC, n = 182), Ganly *et al*. (Hurthle cell thyroid cancer [HTC], n = 56), Gopal *et al*. (HTC, n = 32) and Yoo *et al*. (ATC, n = 27; PDTC, n = 15).

**Supplementary Table 10: Overlap of somatic SNVs across tumor components**

Twenty-eight patients (19 by whole exome sequencing [WXS], 9 by whole genome sequencing [WGS]) had SNV calls for multiple tumor regions (ATC, co-occurring differentiated thyroid cancer [DTC], metastasis or recurrent tumor); hypergeometric tests were used to determine whether there was more overlap than expected by chance alone across regions. A single pair (primary ATC and cervical metastasis) demonstrated no overlap in SNV profile.

**Supplementary Table 11: Overlap of CNAs across tumor components**

Twenty-one patients had copy number aberrations (CNA) calls for multiple tumor regions (ATC, co-occurring DTC, metastasis or recurrent tumor) based on CNA arrays; hypergeometric tests were used to determine whether there was more overlap than expected by chance alone across regions. There was frequently less overlap than expected.

## Notes

### Competing Interest Statement

The authors have declared no competing interest.

### Author Declarations

The study was approved by the Research Ethics Boards at Western University (REB 7182) and informed consent was obtained from each patient.

### Summary of Updates

Abstract and title updated

